# Association between oral health and swallowing function

**DOI:** 10.1101/2022.09.13.22279920

**Authors:** Takafumi Yamano, Kensuke Nishi, Fumitaka Omori, Kaori Wada, Masahiro Yamaguchi, Toru Naito

## Abstract

**Objective:** Oral care is effective for preventing pneumonia, but objective evaluation of its effect on swallowing is lacking. Oral assessment scores, such as the Oral Health Assessment Tool (OHAT), are used to evaluate the oral environment to monitor oral care. Methods: We investigated the relationship between OHAT score and swallowing function in 24 patients aged 64–97 years.

**Results:** The OHAT score did not correlate with the video endoscopy (VE) or video fluorography (VF) scores. Furthermore, the OHAT score was not significantly different between the multiple and no or single pneumonia episode groups. The group with multiple episodes of pneumonia had lower VE and VF scores than that with no or a single episode of pneumonia (p < 0.01).

**Conclusions:** Oral assessment, VE, and VF scores are necessary to evaluate swallowing in patients with suspected dysphagia. In addition, recurrent pneumonia among patients with a good oral environment has multiple contributing factors, including subclinical aspiration, pharyngeal clearance, and delayed activation of the gag reflex.

## 1. Introduction

Although several studies have reported that oral care prevents pneumonia [1–3], few have evaluated the relationship between the oral environment and swallowing function. Anecdotal evidence suggests that oral care has limited efficacy for preventing aspiration and gastroesophageal reflux, especially in older patients.

Regular assessments and standardized oral care protocols are important to provide high-quality oral care that does not vary between caregivers [4]. The Oral Health Assessment Tool (OHAT), among other scales, is used in clinical practice to objectively assess the oral environment.

In the present study, we investigated the relationship of OHAT with swallowing function, assessed using video endoscopy (VE) and video fluorography (VF).

## 2. Materials and Methods

### 2.1 Patients

All patients admitted to our hospital receive a basic oral health check, including OHAT, by a dental hygienist to identify patients who require dental treatment and oral care. Patients with swallowing dysfunction are referred to the Ear, Nose, and Throat department for VE and VF, followed by treatment and rehabilitation.

The study included 24 patients (7 males and 17 females; age range: 64–97 years; average age: 86 years) who underwent OHAT, VE, and VF at Fukuoka Dental College Hospital between April 2014 and October 2019. We excluded patients with head and neck cancer from the study because it may affect the oral environment. The diagnosis of pneumonia was made in cases fulfilling the following criteria (1) and (2). (1) Chest X-ray or chest computed tomography showing alveolar infiltration shadow. (2) Fever of 37.5°C or higher and abnormally high levels of C-reactive protein. Peripheral white blood cell count of more than 9000/μL. Presence of any two or more airway symptoms such as sputum. We evaluated the associations of OHAT with VE and VF, and compared the associations between patients with no, or a single episode, of pneumonia and patients with multiple episodes.

### 2.2 Evaluation method

OHAT was performed to evaluate the oral environment [5]. Scores for items pertaining to the lips, tongue, gingival mucosa, saliva, remaining teeth, dentures, oral cleaning, and toothache were recorded on a scoring sheet using a three-point scale (0–2). Higher OHAT scores indicate a worse oral environment.

VE was evaluated using the Hyodo scoring system [6] based on four parameters: salivary pooling at the vallecula and piriform sinuses; induction of glottal closure reflex on touching the epiglottis or arytenoid with the endoscope; swallowing reflex initiation, assessed based on the “whiteout” timing (where whiteout was defined as the period during which the endoscopic image is obscured due to pharyngeal closure); and pharyngeal clearance after swallowing blue-dyed water. These parameters were scored on a 4-point scale (0: normal; 1: mildly impaired; 2: moderately impaired; 3: severely impaired), with higher scores indicating greater swallowing dysfunction.

VF was evaluated using a method described previously [7]. The patient was placed in a sitting or semi-sitting position, and asked to swallow 10 cc of non-ionic iodinated contrast agent (Omnipaque 300^®^) for a single examination. Swallowing was classified as normal, < 50% dysphagia, and > 50% dysphagia. Early pharyngeal (present or absent) and laryngeal (none, only laryngeal inflow, or laryngeal inflow with aspiration) inflows were also recorded. Laryngeal elevation delay time (LEDT) is a useful visual marker of the onset of pharyngeal swallowing; an LEDT ≤ 0.35 s is considered normal [8]. Prolongation was categorized as no residual epiglottic trough with minor prolongation; no overflow from the epiglottic trough with overflow into the pharynx; no residual pear-shaped depression with minor prolongation and no overflow into the larynx; and overflow into the larynx.

### 2.3 Data analysis

Spearman’s rank correlation coefficient and Welch’s t-test were used to analyze the data. Data are expressed as mean ± standard error of the mean (SEM).

The study was approved by the Fukuoka Gakuen Research Ethics Committee (permission no. 314).

## 3. Results

### 3.1 Primary diseases and oral intake

The most common disease was pneumonia (17 patients), followed by cerebral infarction (5 patients), pyelonephritis (4 patients), bronchitis (2 patients), Parkinson’s disease (2 patients), scleroderma (1 patient), diabetes (1 patient), esophageal cancer (1 patient), and Parkinson’s syndrome (1 patient). Some patients had multiple diseases. Oral intake was possible in 20 patients (80%), whereas tube feeding and gastric banding were required in 4 patients.

### 3.2 Correlation between OHAT and VE

No correlation was observed between the OHAT and VE scores (r = 0.3153; Fig. 1A). In Figure 1A, the vertical and horizontal axes represent the VE and OHAT scores, respectively.

**Figure 1A.**
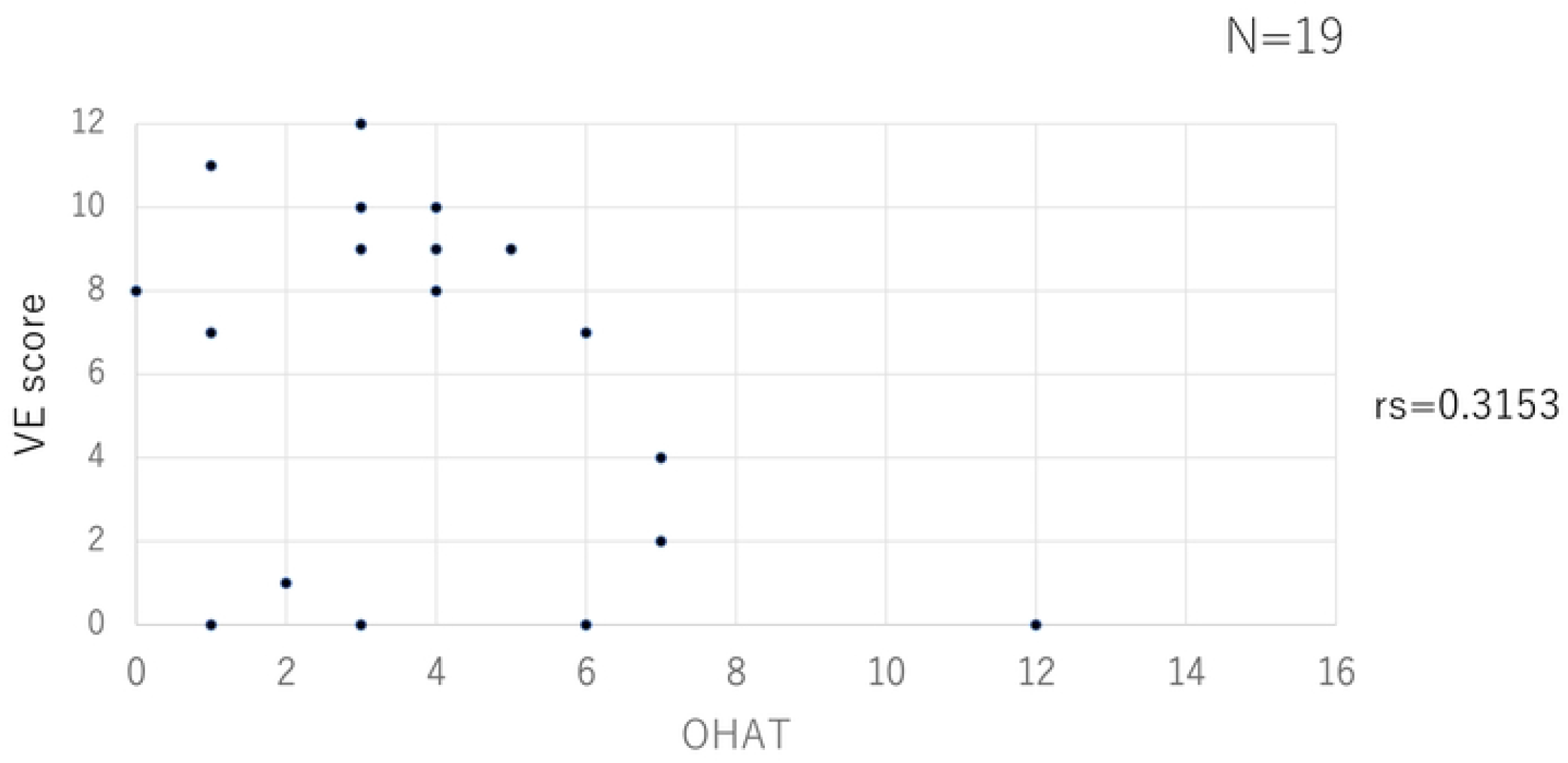
There was no correlation between the Oral Health Assessment Tool (OHAT) and video endoscopy (VE) scores (r = 0.2211).

### 3.3 Correlation between OHAT and VF scores

No correlation was observed between the OHAT and VF scores (r = 0.2211; Fig. 1B).

**Figure 1B.**
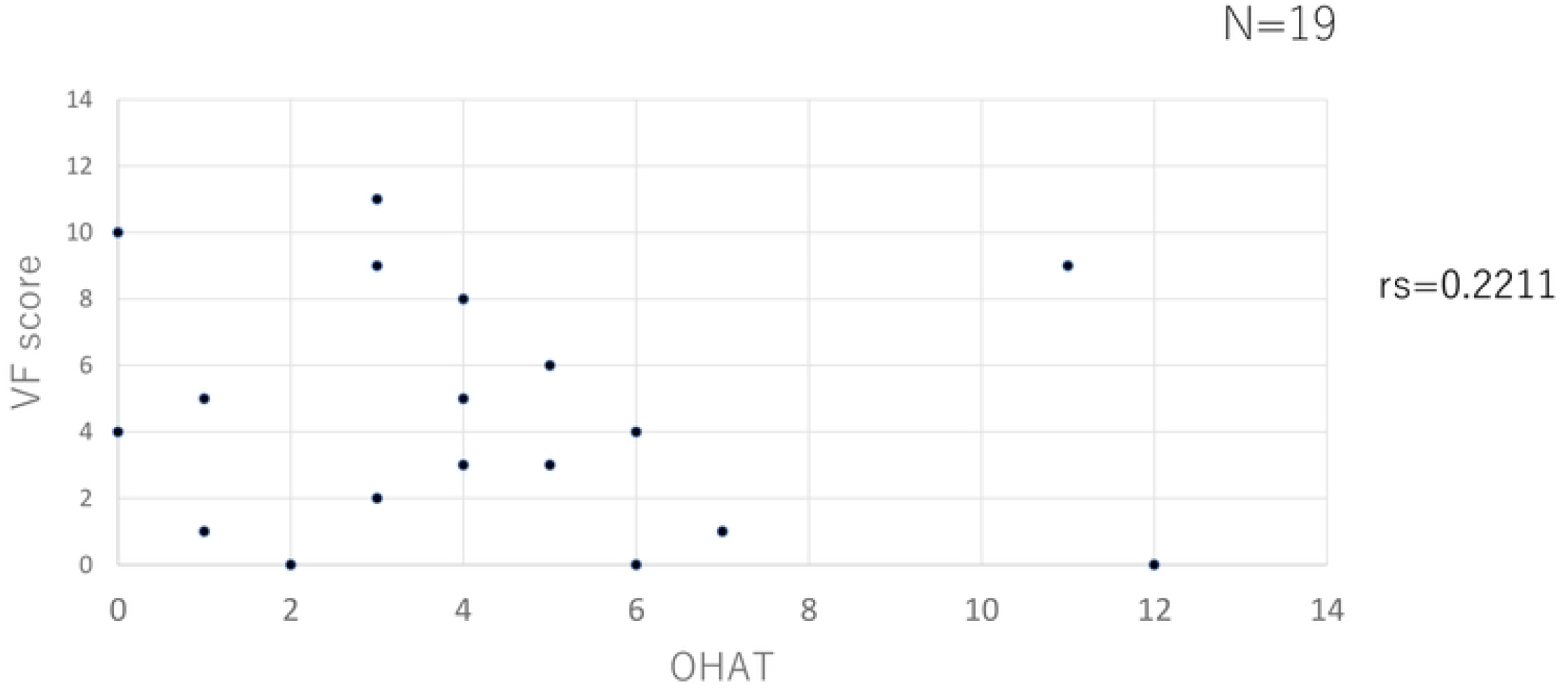
There was no correlation between the Oral Health Assessment Tool (OHAT) and video fluorography (VF) scores (r = 0.2211).

### 3.4 Comparison between the no or single and multiple pneumonia episode groups

No significant difference was observed in the OHAT score between the no or single and multiple pneumonia episode groups (Figure 2).

**Figure 2.**
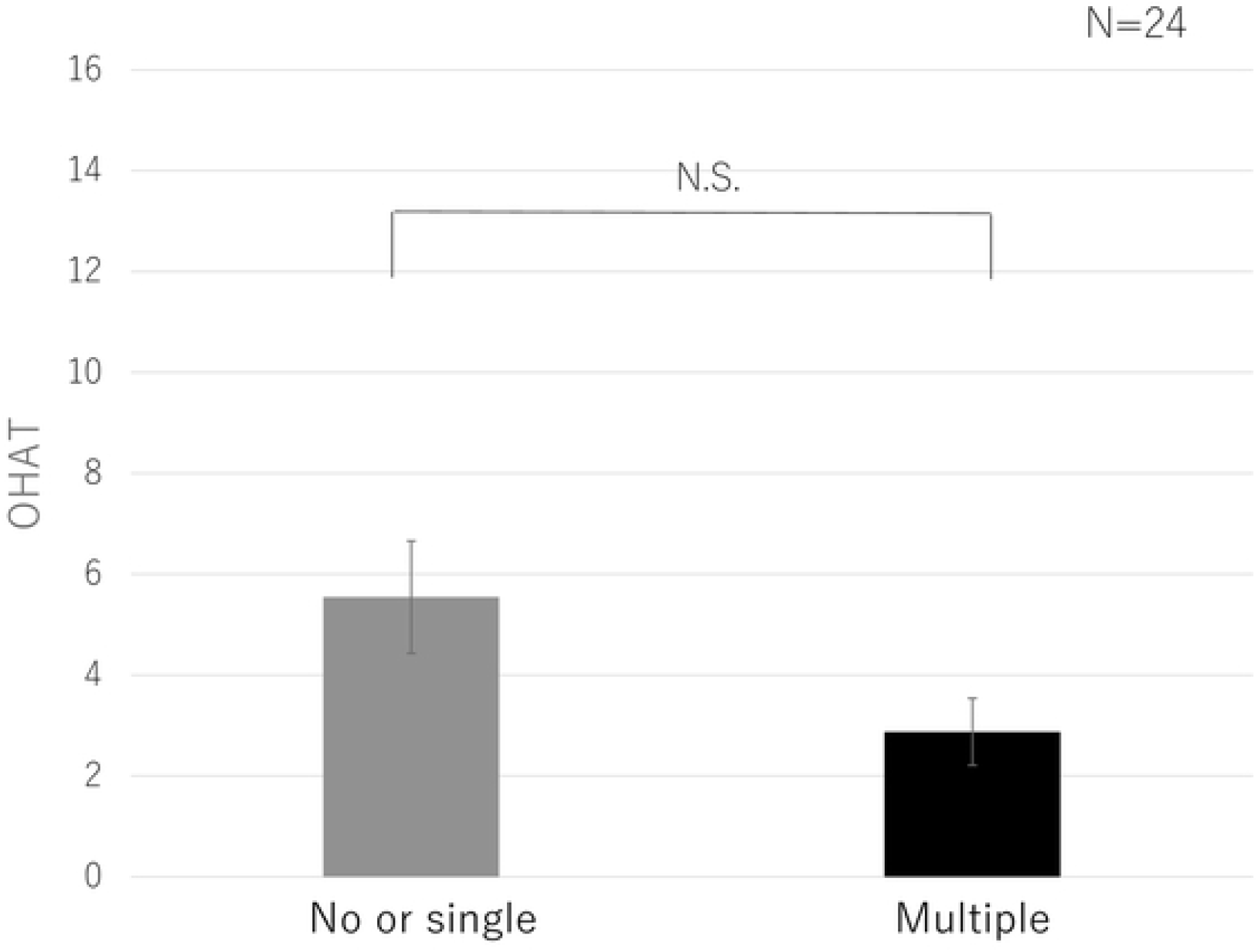
Comparison of the Oral Health Assessment Tool (OHAT) score between the no or single and multiple pneumonia episode groups.

The VE score was significantly higher in the multiple compared to no or single pneumonia episode group (p < 0.01; Fig. 3A). Analysis of the OHAT items showed that scores for salivary retention in the laryngeal trough and pisiform depression, elicitation of glottic closure, cough, and gag reflexes, and pharyngeal clearance by colored water swallow were higher in the multiple compared to no or single pneumonia episode group (Fig. 3B).

**Figure 3A.**
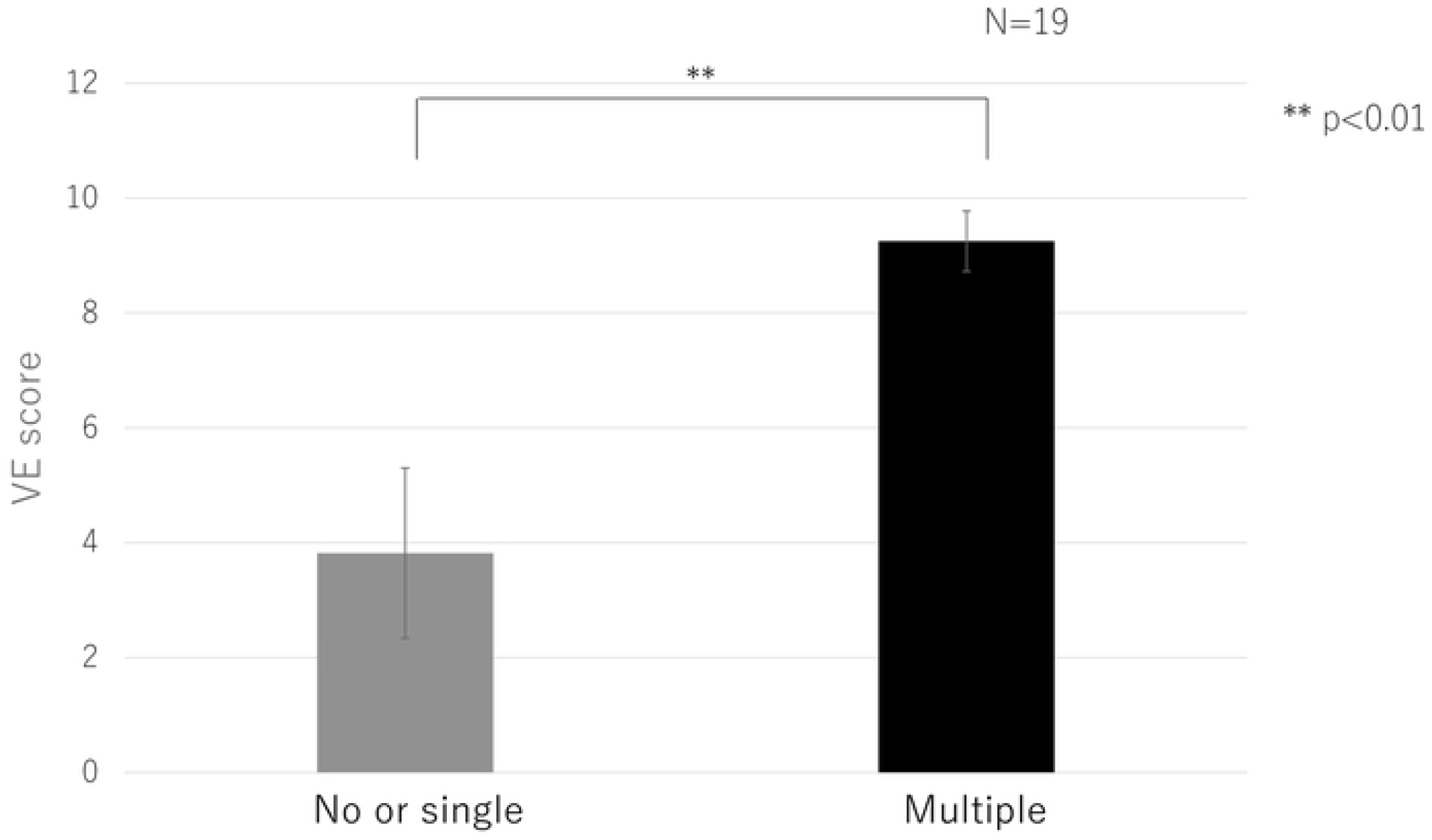
Comparison of the video endoscopy (VE) score between the no or single and multiple pneumonia episode groups.

**Figure 3B.**
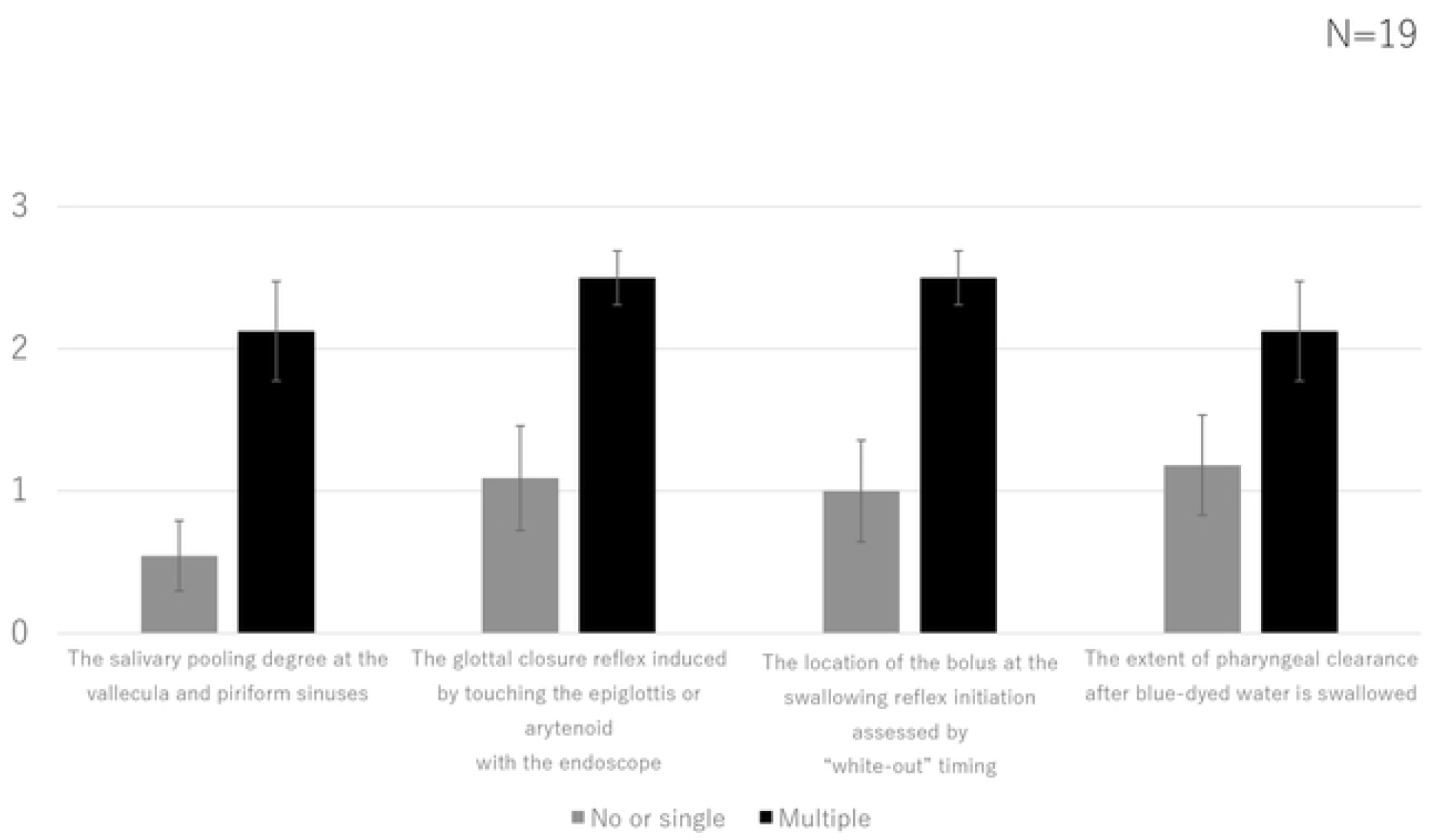
Comparison of the video endoscopy (VE) item scores between the no or single and multiple pneumonia episode groups.

Similar to the VF score, the swallowing angiography score was significantly higher in the multiple compared to no or single pneumonia episode group (p < 0.01; Fig. 4A). Patients with multiple episodes of pneumonia had higher scores for all items compared to those with no or a single episode of pneumonia (Fig. 4B).

**Figure 4A.**
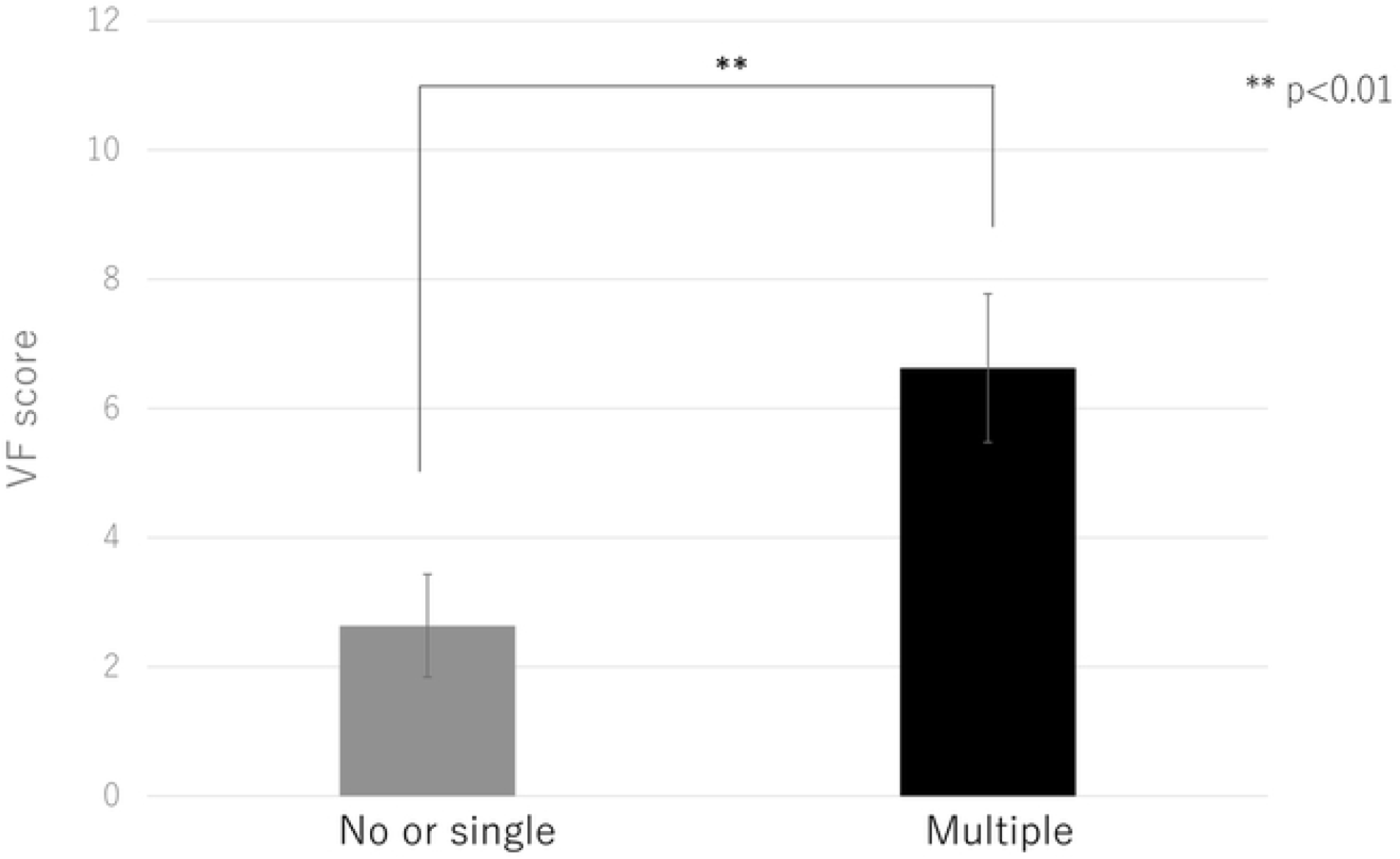
Comparison of the video fluorography (VF) score between the no or single episode and multiple pneumonia episode groups.

**Figure 4B.**
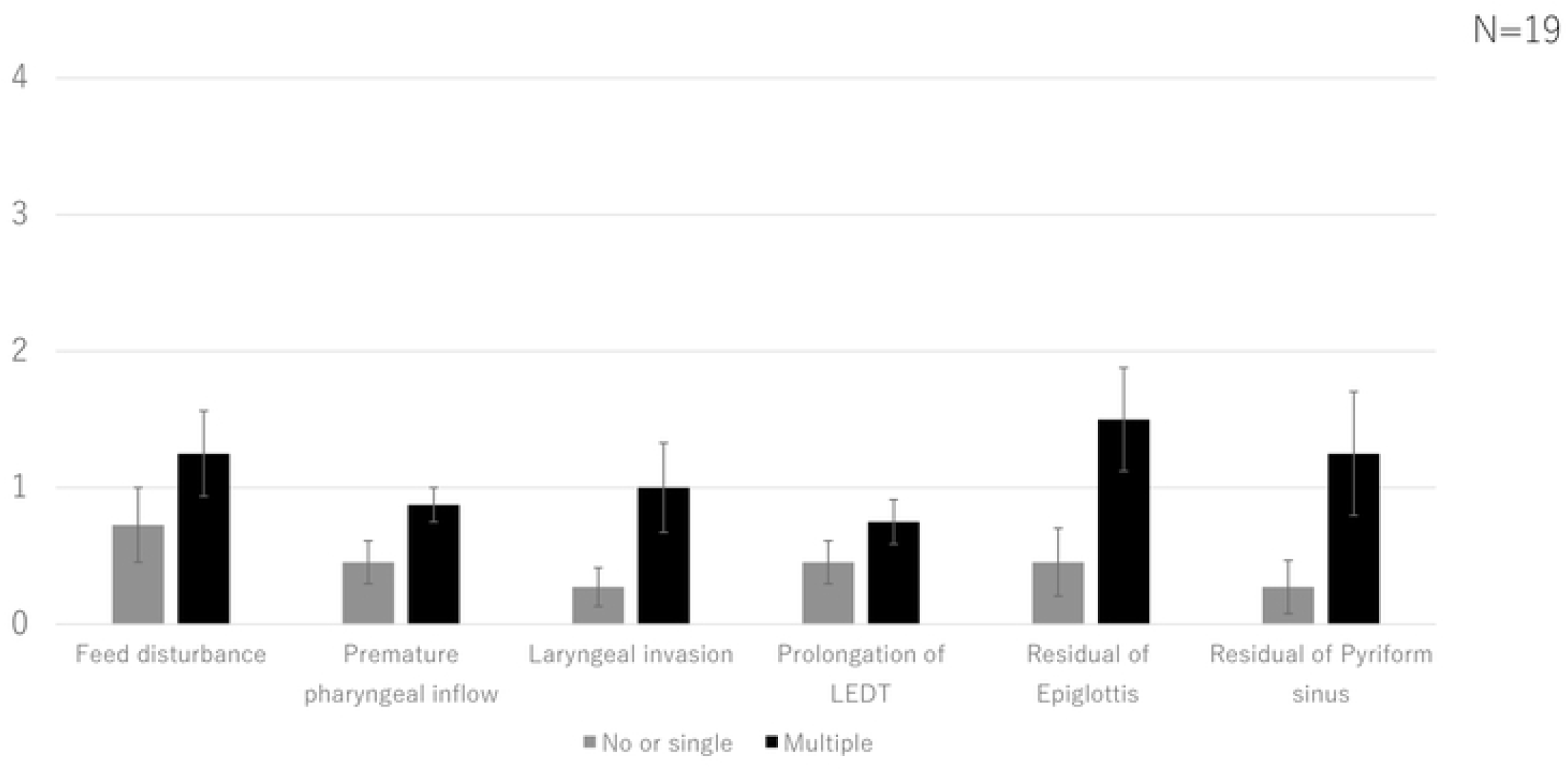
Comparison of the video fluorography (VF) item scores between the no or single and multiple pneumonia episode groups.

## 4. Discussion

Some previous studies have suggested that oral care prevents pneumonia in elderly patients, with a relative risk of pneumonia of 19% and 11% in elderly patients with poor and good oral health, respectively. In addition, previous studies reported that elderly patients with poor oral health had a 1.7-fold higher relative risk of pneumonia compared to those with good oral health [1]. Past reports also suggest that the cough reflex threshold was significantly reduced after 1 month of meticulous oral care in a nursing home for the elderly compared to that before the start of care [2]. Furthermore, individuals receiving oral care from a dental hygienist twice a week for 24 months in nursing homes had a significantly lower likelihood of fever and deaths from aspiration pneumonia compared to individuals who did not receive oral care [3]. Therefore, oral care is recommended as part of the treatment of dysphagia, especially in elderly residents of nursing homes.

Although the oral environment significantly affects the risk of pneumonia, few studies have evaluated its relationship with swallowing. Thus, there is no doubt that there is a significant association between the oral environment and the development of pneumonia, but there are few reports comparing VE andVF, which reflect actual swallowing function. Nakayama et al. [9] studied patients admitted to a convalescent hospital and found that the Functional Oral Intake Scale (FOIS) score was significantly associated with the saliva and denture scores. However, after excluding patients on non-oral nutrition, no significant association between FOIS and OHAT scores was seen.

Importantly, the FOIS score reflects oral intake status rather than swallowing function. Recent studies using the Hyodo scoring system have shown that VE is simple to perform and effective for predicting aspiration, with a score of ≥ 6 corresponding to the highest risk of aspiration [11]. The Hyodo score significantly correlates with handgrip strength and peak expiratory flow rate, making it useful for examining the effects of strength training on dysphagia [12].

VF is the most reliable swallowing assessment method because it can evaluate the oral, pharyngeal, and esophageal phases [13]. In addition, VF is extremely useful when making policy decisions. In VF, whiteout due to pharyngeal contraction does not occur and the oral and esophageal phases, which cannot be observed with VE, can be evaluated. At our institution, VF is also used to determine the timing of a return to of oral intake after prolonged fasting, and the optimal treatment for swallowing rehabilitation or surgery to improve swallowing.

In the present study, we did not identify any correlations between the OHAT score for oral assessment and VE and VF scores for swallowing. In addition, the OHAT score did not differ between the no or single and multiple pneumonia episode groups, while the rates of swallowing endoscopy and angiography were significantly different between the groups. In our previous study [9], we identified significant differences in the oral and pharyngeal phases on VE between elderly patients with pneumonia and those without a history of pneumonia; there was a particularly significant group difference in elicitation of the gag reflex. These findings suggest that for patients with fever, difficulty in oral intake, or other findings suggestive of dysphagia, VE or VF for the evaluation of swallowing in addition to oral assessment are necessary. This suggests that the main cause of pneumonia in the elderly is thought to occur when both poor oral and poor pharyngeal swallowing function are present, but it was speculated that the greater factor is poor pharyngeal swallowing function. However, the present study is very limited, as it is not simply a comparison of patients with and without a history of aspiration pneumonia.

Age-related swallowing dysfunction is multifactorial, with 63% of elderly patients (mean age: 83 years) having oral abnormalities, such as difficulty ingesting, controlling, or delivering a bolus to initiate swallowing; this prevalence rate was higher compared to young patients without dysphagia. In addition, 25% of the elderly patients had pharyngeal dysfunction, such as bolus retention and tongue propulsion or pharyngeal muscle paralysis, and 36% had esophageal abnormalities due to cricopharyngeal muscle dysfunction [14]. Age-related swallowing dysfunctions in the elderly without organic diseases, such as stroke, cancer, and dementia, include prolonged oral transit time and aspiration [15].

In the present study, the multiple episodes of pneumonia group had higher VE and VF scores for all items compared to the no or single episode of pneumonia group. Thus, multiple factors, including subclinical aspiration, pharyngeal clearance, and delayed activation of the gag reflex, likely promote recurrent pneumonia among patients with a good oral environment.

The authors declare no conflicts of interest associated with this manuscript.

The datasets generated and/or analyzed during the current study are available from the corresponding author on reasonable request.

## Data Availability

Ethics Committee (contact via the Fukuoka Gakuen Research Ethics Committee) for researchers who meet the criteria for access to confidential data.

## Acknowledgments

None

---

**Table 1.**
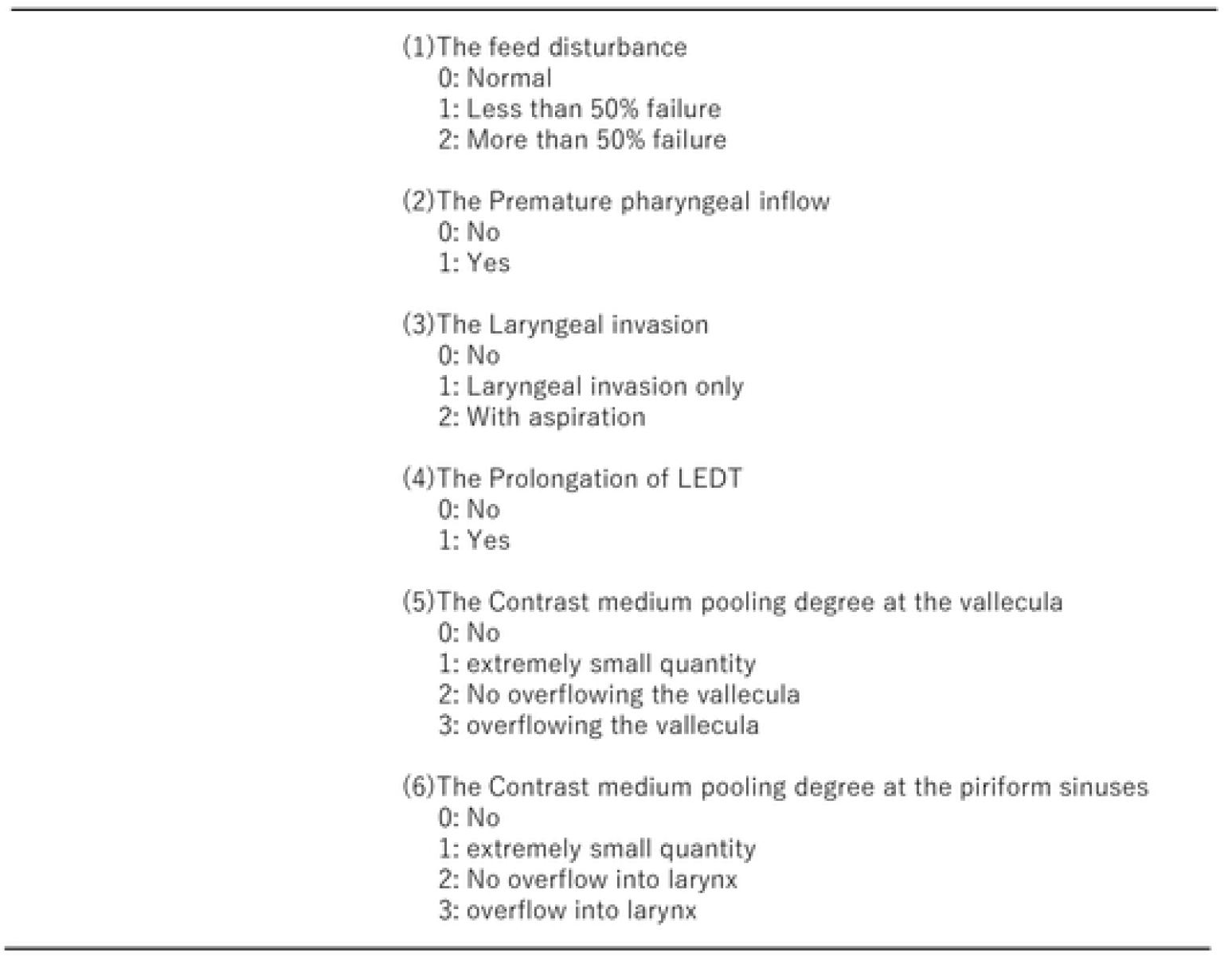
Video fluorography (VF) scoring system proposed by Yamano et al. in 2019 [7]. The English in this document has been checked by at least two professional editors, both native speakers of English. For a certificate, please see: http://www.textcheck.com/certificate/0JN9dx

